# Core Outcome Set for Clinical Trials of COVID-19 based on Traditional Chinese and Western Medicine

**DOI:** 10.1101/2020.03.23.20041533

**Authors:** Ruijin Qiu, Chen Zhao, Tengxiao Liang, Xuezeng Hao, Ya Huang, Xiaoyu Zhang, Zhao Chen, Xuxu Wei, Mengzhu Zhao, Changming Zhong, Jiayuan Hu, Min Li, Songjie Han, Tianmai He, Jing Chen, Hongcai Shang

**Author notes:** **Corresponding author:** 1. Prof. Jing Chen, Postal address: 88th Yuquan Road, Nankai District, Tianjin, China, 300193, 2. Prof. Hongcai Shang, Postal address: Hai Yun Cang on the 5th Zip, Dongcheng District, Beijing, China, 100700.

## Abstract

**Background:** Development of a core outcome set (COS) for clinical trials for COVID-19 is urgent because of the pandemic wreaking havoc worldwide and the heterogeneity of outcomes in clinical trials.

**Methods:** A preliminary list of outcomes were developed after a systematic review of protocols of clinical trials for COVID-19. Then, two rounds of the Delphi survey were conducted. Stakeholders were traditional Chinese medicine (TCM) experts, Western medicine (WM) experts, nurses and the public. Patients with confirmed COVID-19 were also invited to participate in a questionnaire written in understandable language. Frontline clinicians, as well as nurse, methodologist, evidence based-medicine researcher, and staff from the Chinese Clinical Trials Registry participated by video conference to vote.

**Results:** Ninety-seven eligible study protocols were identified from 160 clinical trials. Seventy-six outcomes were identified from TCM clinical trials and 126 outcomes were identified from WM clinical trials. Finally, 145 outcomes were included in the first round of the Delphi survey. Then, a COS for clinical trials of TCM and WM was developed. The COS include clinical outcomes (recovery/improvement/progression/death), etiology (SARS-CoV-2 nucleic-acid tests, viral load), inflammatory factor (C-reactive protein), vital signs (temperature, respiration), blood and lymphatic-system parameters (lymphocytes, virus antibody), respiratory outcomes (Pulmonary imaging, blood oxygen saturation, PaO2/FiO2 ratio, arterial blood gas analysis, mechanical ventilation, oxygen intake, pneumonia severity index), clinical efficacy (prevalence of preventing patients with mild-to-moderate disease progressing to severe disease), symptoms (clinical symptom score). Outcomes were recommended according to different types of disease. Outcome measurement instrument/definition were also recommended.

**Conclusion:** A COS for COVID-19 may improve consistency of outcome reporting in clinical trials.

## 1. INTRODUCTION

After a novel coronavirus causing pneumonia was spread in since December 2019 around the world, it was named temporarily as “2019 novel coronavirus” on 2 January 2020. Then the World Health Organization (WHO) officially named it as “coronavirus disease 2020” (COVID-19) on 11 February 2020. The coronavirus that causes COVID-19 was termed “severe acute respiratory syndrome coronavirus 2” (SARS-CoV-2) on 11 February 2020 by the WHO.

As of midnight on 18 March 2020, 191,127 cases of COVID-19 have been reported to the WHO, and 7,807 of them have died [1]. At the same time in China, according to the Internet website of the National Health Commission of the People’s Republic of China, 80,928 confirmed cases have been reported from all areas of China [2].

COVID-19 is now a global threat, so its outbreak was declared to be a pandemic on 11 March 2020 by the WHO. There are significant knowledge gaps in the epidemiology, transmission dynamics, investigation tools, and management of COVID-19 [3]. A specific drug or vaccine has not been approved to treat it. Hence, COVID-19 management is a major challenge for clinicians and researchers worldwide.

The first clinical trial of COVID-19 was registered on 23 January 2020 [4]. Since then, an increasing number of clinical trials of COVID-19 have been registered using regimens based on traditional Chinese medicine (TCM) and Western medicine (WM). As of March 18, 2020, 585 protocols were searched from all the databases of International Committee of Medical Journal Editors (ICMJE)-accepted platforms of clinical-trial registries.

Previously, we found several problems regarding the protocols of clinical trials of COVID-19 (e.g., unclear study objectives, heterogeneity of outcome choices, and small study population [5]) that may reduce the value of clinical trials. In the meantime, clinicians’ understanding of COVID-19 characteristics has been changing because they are treating many more patients than before. The diagnosis and management plan of COVID-2019 also keeps changing. We believe that certain inappropriate outcomes may be chosen by researchers. To improve the consistency of outcomes and include more clinical trials in systematic reviews, development of a “core outcome set” (COS) for COVID-19 is crucial.

A COS is an agreed standardized set of outcomes that should be measured and reported, as a minimum, in all clinical trials in specific areas of health or healthcare [6]. When researchers report outcomes in a COS, they can also report other outcomes.

This COS was based on: (i) a population with confirmed COVID-19 of “mild”, “ordinary”, “severe” or “critical” types; (ii) interventions that include TCM and WM; (iii) the COS being applied in randomized controlled trials (RCTs) and observational studies.

## 2. METHODS

### 2.1 Registry

This COS has been registered on the Core Outcome Measures in Effectiveness Trials (COMET) database [7]. This research was conducted and reported following COS-STAndards for Development (COS-STAD) [8] and COS-STAndards for Reporting (COS-STAR) [9].

### 2.2 Participants

#### 2.2.1 Steering group

A steering group was formed by a TCM expert, WM expert, methodologist, nurse and statistician. They conducted the research protocol, made decisions if there was confusion, and attended the consensus meeting to facilitate COS development.

#### 2.2.2 Stakeholders in the Delphi survey

The stakeholders in the Delphi survey included TCM experts (clinicians and researchers), WM experts (clinicians and researchers), nurses, patients and the public.

COVID-19 is a new infectious disease that is spreading rapidly. In China, many clinicians have been trained to face emergencies, irrespective of whether they are on the “frontline” of the “battle” against COVID-19. More than 40,000 clinicians and nurses from other areas of China moved to Hubei Province to support the local medical system. Not all of these clinicians and nurses were trained in respiratory medicine or critical care. To obtain perspectives on a larger scale, we used “snowball” sampling to extend the sample size. We invited members from the Clinical Research Information Association of China, and the Information Association for Traditional Chinese Medicine and Pharmacy to participate in the Delphi survey. We asked them to send the questionnaire to their colleagues.

We believe that the perspectives of patients and public are important. Hence, we sent the questionnaire via social media (WeChat, Tencent) to invite the public to participate.

To obtain patients’ perspectives, frontline clinicians of Dongzhimen Hospital within Beijing University of Chinese Medicine (Beijing, China) invited and helped patients who consented to complete the questionnaire.

#### 2.2.3 Stakeholders in the consensus meeting

The stakeholders in the consensus meeting were a TCM clinician, WM clinician, nurse, methodologist, evidence-based medicine researcher and staff from the Chinese Clinical Trials Registry.

### 2.3 Information Sources

All the databases of ICMJE-accepted platforms of clinical-trial registries [10] were considered. Search terms for Chinese Clinical Trial Registry (ChiCTR) were: “COVID-19,” “2019-novel Corona Virus (2019-nCoV),” “Novel Coronavirus Pneumonia (NCP),” “Severe Acute Respiratory Infection (SARI),” and “Severe Acute Respiratory Syndrome - Corona Virus-2 (SARS-CoV-2).” Search terms for the Netherlands National Trial Register were “nCoV,” “Coronavirus,” “SARS,” “SARI,” “NCP,” and “COVID.” Search terms for other databases were “2019-nCoV OR Novel Coronavirus OR New Coronavirus OR SARS-CoV-2 OR SARI OR NCP OR Novel Coronavirus Pneumonia OR COVID-19 OR Wuhan pneumonia.”

The search was conducted on 14 February 2020. The details of inclusion criteria, exclusion criteria, study identification, date extraction, rejected/combined outcomes are described in the systematic review of protocols of clinical trials of COVID-19 [11].

### 2.4 Consensus Process

Two rounds of the Delphi survey for professionals and the public, as well as one round of the Delphi survey for patients, were conducted. After the Delphi survey had been completed, a consensus meeting was conducted to determine the final COS.

#### 2.4.1 Delphi survey

The questionnaire for professionals and the public was sent by smartphone. It included individual outcome in different outcome domains and scoring. At the end of the questionnaire, there were two open-ended questions: (i) which outcomes do you think are important but which were not included in the questionnaire? (ii) what is your opinion of this questionnaire?

The questionnaire for patients was sent by smartphone, too. It included outcomes/outcome domains that were understood readily by patients. Patients were asked to vote which outcomes/outcome domains were important to them. There was one open-ended question: which outcomes do you think are important but which were not included in the questionnaire?

#### 2.4.2 Outcome scoring

The questionnaire for professionals and the public employed a nine-point scoring system, which has been used in a previous COS [12, 13]. A score of: “1–3” denoted that the outcome was not important for inclusion in the COS; “4–6” meant the outcome was important but not critical for inclusion in the COS; “7–9” denoted that the outcome was critical for inclusion in the COS. An outcome scored as <7 by ≤50% of participants for all stakeholders was removed from the next consensus process. The outcomes recommended by participants were added in the second round of the Delphi survey after discussion by the steering group.

#### 2.4.3 Consensus definitions

For the Delphi survey administered to professionals and the public, the consensus definitions were: (i) consensus in: ≥70% of participants in all stakeholders scored the outcome as 7–9, and <15% of participants in all stakeholders scored the outcome as 1–3; (ii) consensus out: ≤50% of participants in TCM experts and WM experts scored the outcome as 7–9; (iii). no consensus: anything else.

The voice of patients should be considered. Hence, for the patients’ survey, the consensus definition was outcomes that were voted by >50% of patients. For the consensus meeting, the consensus definitions were: (i) consensus in: outcomes that were voted by ≥70% of participants; (ii). consensus out: outcomes that were voted by <70% of participants.

#### 2.4.4 Consensus meeting

The consensus meeting was held by teleconference. The contents of the consensus meeting covered: (i) the reporting background and methods of the research; (ii) reporting the results of the Delphi survey of professionals and the public, and the results of the patients’ questionnaire; (iii) discussing the candidate outcomes and their instruments/definition; (iv) voting on the outcomes and reaching a consensus.

### 2.5 Ethics and consent

The entire project is part of a clinical trial of COVID-19, which was approved by the Ethics Committee of Dongzhimen Hospital (DZMEC-KY-2020-09). Because of the special circumstances of the COVID-19 pandemic, participants who completed the questionnaire were assumed to have provided consent for their data to be used.

## 3. RESULTS

A total of 160 protocols from 19 platforms of clinical-trial registries were searched. After reading the titles and study details, 63 non-relevant or ineligible study protocols were excluded. Finally, 97 eligible study protocols were included from ChiCTR and ClinicalTrials.gov. Thirty-four clinical trials were for TCM therapy and 63 clinical trials were for WM therapy. All clinical trials will be conducted in China. These clinical trials comprised 75 RCTs (53 for WM and 22 for TCM) and 22 non-RCTs (10 for WM and 12 for TCM). For 34 protocols of TCM clinical trials, there were 76 individual outcomes from 16 outcome domains after the merging and grouping of outcomes. For 63 protocols of WM clinical trials, there were 126 individual outcomes from 17 outcome domains after merging and grouping. The list of outcomes can be obtained from [11]. There were >40 duplicated outcomes between the TCM and WM protocols for clinical trials.

After removing duplicated outcomes, we developed the first round of the Delphi survey. After review by the steering group, 145 outcomes were included in the questionnaire.

### 3.1 Round 1 of the Delphi survey

We had incentive measures to improve the response of the Delphi survey (randomized rewards after completing and submitting the questionnaire). The time planned for round 1 of the Delphi survey was from 4 March to 12 March 2020. As of March 9, 2020, 176 participants had completed the questionnaire. After review, 51 questionnaires were found to be invalid. On March 8 and 9, 2020, ≤ 5 questionnaire/day were completed, and almost all of them were invalid. Most of the invalid questionnaires had been completed by the public within 5 min (it was not possible for people who were unfamiliar with COVID-19 to complete the questionnaire) or who had chosen the same score for all outcomes. After discussion with the steering group, we decided to stop the Delphi survey.

Finally, 125 valid questionnaires were evaluated. The characteristics of participants in the round 1 of Delphi survey are shown in Table 1. The number of outcomes that achieved consensus and no consensus in different stakeholders are shown in Table 2. The list of outcomes is shown in Supplement 1.

**Table 1.**
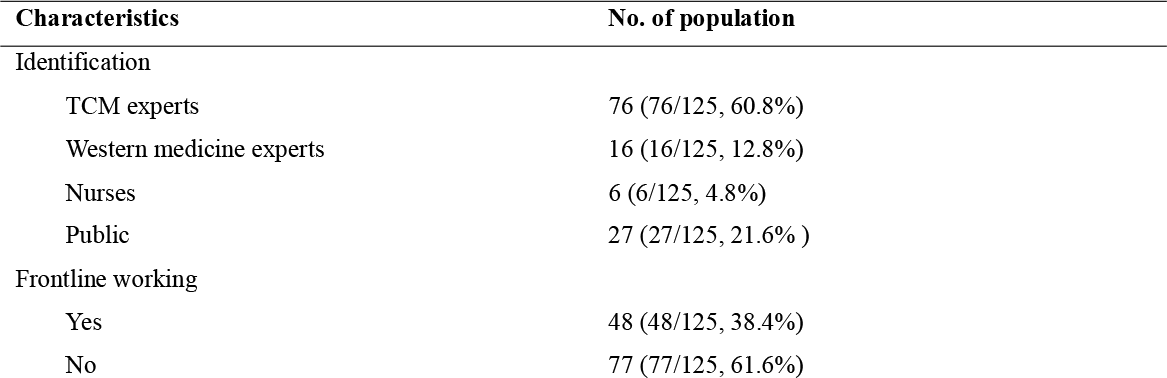
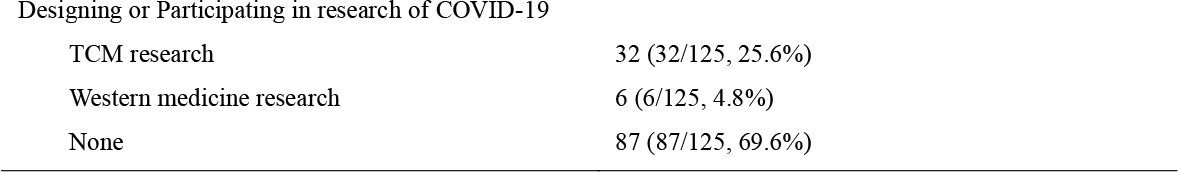
**The characteristic of participants in the round 1 of Delphi survey**

| Characteristics | No. of population |
| --- | --- |
| Identification |  |
| TCM experts | 76 (76/125, 60.8%) |
| Western medicine experts | 16 (16/125, 12.8%) |
| Nurses | 6 (6/125, 4.8%) |
| Public | 27 (27/125, 21.6% ) |
| Frontline working |  |
| Yes | 48 (48/125, 38.4%) |
| No | 77 (77/125, 61.6%) |
| Designing or Participating in research of COVID-19 |  |
| TCM research | 32 (32/125, 25.6%) |
| Western medicine research | 6 (6/125, 4.8%) |
| None | 87 (87/125, 69.6%) |

**Table 2.** The number of outcomes that achieved consensus and no consensus in round 1 of Delphi survey

| Stakeholders | Consensus in | Consensus out | No consensus |
| --- | --- | --- | --- |
| TCM experts | 34 | 50 | 61 |
| Western medicine experts | 50 | 47 | 48 |
| Nurses | 126 | 2 | 17 |
| Public | 106 | 0 | 39 |

Only 15 (15/125, 12%) participants were in Hubei Province. However, the Internet Protocol (IP) address that the electronic questionnaire obtained showed that 30 (30/125, 24%) participants were in Hubei Province. The regions of participants are shown in Figure 1.

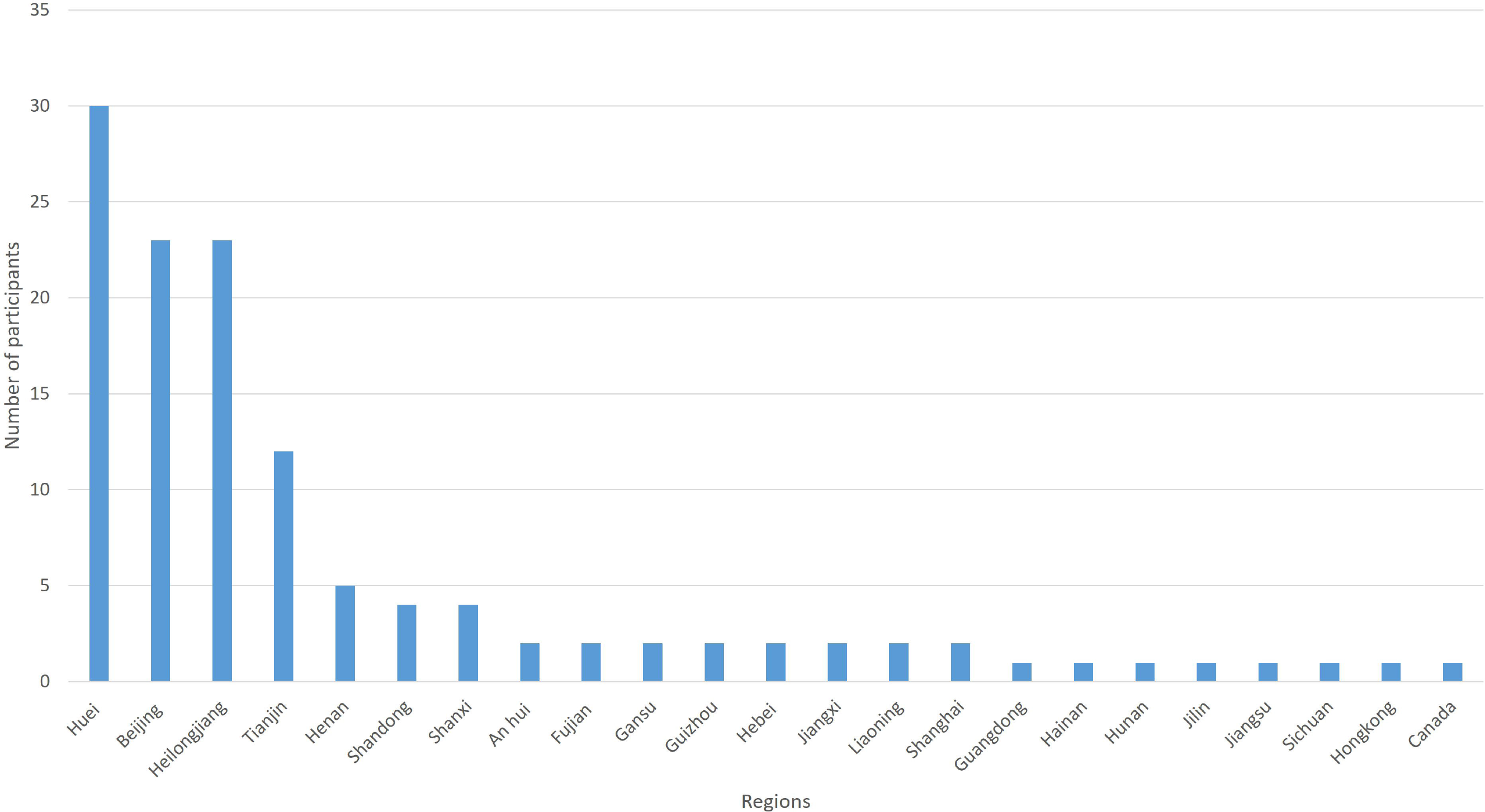

More than 20 participants provided outcomes or significant proposals for round 1 of the Delphi survey. After discussion with the steering group, six of them were added to round 2 of the Delphi survey. No “consensus out” outcomes by all of the stakeholders were noted. The steering group discussed the results of round 1 of the Delphi survey. The steering group believed that for nurses and the public, it was too difficult to give-up some outcomes because of the knowledge gap. They decided that all of the outcomes should be entered in round 2 of the Delphi survey.

### 3.2 Round 2 of the Delphi survey

According to the significant proposals from participants in round 1 of the Delphi survey, the steering group decided to add more personal information. To reduce the risk of invalid questionnaires, participants would receive a random reward if the completed questionnaire was considered to be valid. Participants were also asked if they agreed to be mentioned in the “acknowledgements” section when the research was published. Computed tomography and magnetic resonance imaging of the hip were grouped as “hip imaging”. There were 150 individual outcomes in round 2 of the Delphi survey.

The feedback from participants in round 1 of the Delphi survey showed that scoring for some outcomes was difficult. Hence, in round 2 of the Delphi survey, participants had the opportunity to choose “unclear” for any outcome that was difficult to determine. The median score of each outcome from each stakeholder group was shown in round 2 of the Delphi survey. The steering group wanted more people to participate in the Delphi survey. Hence, the questionnaire was sent to potential participants (irrespective of whether they completed round 1 of the Delphi survey) and they were asked to invite colleagues who might be interested in this research. Round 2 of the Delphi survey was conducted from 11 to 13 March 2020.

A total of 110 questionnaires were completed, but seven of them were invalid, so 103 valid questionnaires were assessed. The characteristics of participants in round 2 of the Delphi survey are shown in Table 3.

**Table 3.** The characteristic of participants in the round 1 of Delphi survey

| Characteristics | No. of population | Characteristics | No. of population |
| --- | --- | --- | --- |
| Identification |  | Frontline working |  |
| TCM experts | 60 (60/103, 58.3%) | Yes | 42 (42/103, 40.8%) |
| Western medicine experts | 22 (22/103, 21.4%) | No | 61 (61/103, 59.2%) |
| Nurses | 13 (13/103, 12.6%) | Designing or Participating in research of COVID-19 |  |
| Public | 8 (8/103, 7.8%) | TCM research | 25 (25/103, 24.3%) |
| Education background |  | Western medicine research | 2 (2/103, 1.9%) |
| Doctor | 35 (35/103, 34%) | None | 76 (76/103, 73.8%) |
| Master | 50 (50/103, 48.5%) | Participating in round 1 of Delphi survey |  |
| Undergraduate | 14 (14/103, 13.6%) | Yes | 26 (26/103, 25.2%) |
| Others | 4 (4/103, 3.9%) | No | 77 (77/103, 74.8%) |
| Professional qualification |  |  |  |
| Senior | 30 (30/103, 29.1%) |  |  |
| Intermediate | 46 (46/103, 44.7%) |  |  |
| Junior | 17 (17/103, 16.5%) |  |  |
| None | 10 (10/103, 9.7%) |  |  |

The IP address that the electronic questionnaire obtained showed that 28 (28/103, 27.2%) participants were in Hubei Province. The regions of participants are shown in Figure 2. The number of outcomes that achieved consensus and no consensus in different stakeholders are shown in Table 4. The list of outcomes is shown in Supplement 2.

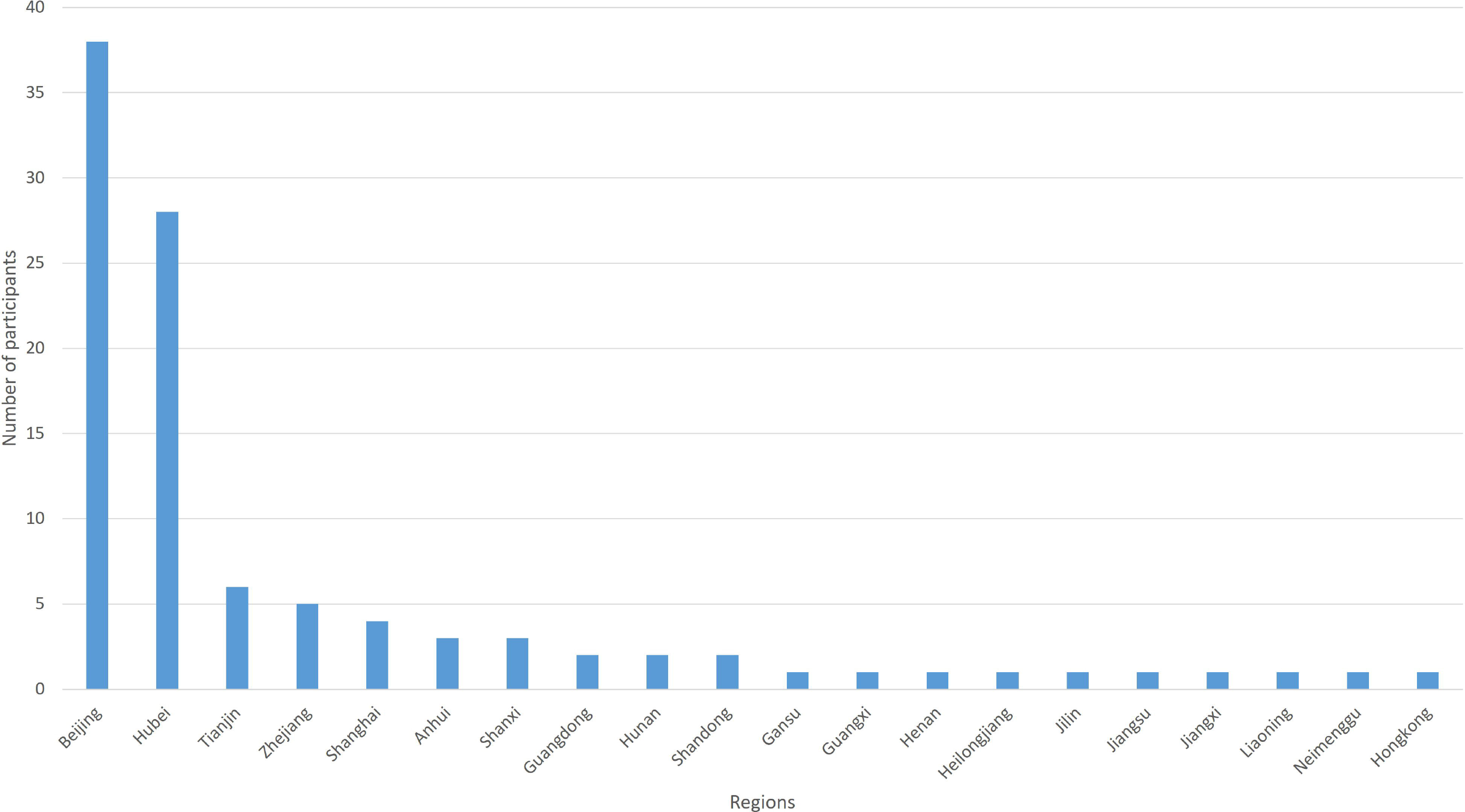

**Table 4.** The number of outcomes that achieved consensus and no consensus in round 2 of the Delphi survey

| Stakeholders | Consensus in | Consensus out | No consensus |
| --- | --- | --- | --- |
| TCM experts | 91 | 35 | 24 |
| Western medicine experts | 57 | 44 | 49 |
| Nurses | 141 | 0 | 9 |
| Public | 104 | 31 | 15 |

After the results of round 2 of the Delphi survey had been reviewed by the steering group, outcomes that achieved “consensus out” by TCM experts and WM experts were excluded. Outcomes that achieved “consensus in” from stakeholders were grouped and presented according to the classification of disease and interventions. They were presented to consensus-meeting participants with “no consensus outcomes” before the consensus meeting was held.

### 3.3 Patients’ survey

Results suggested that nurses and the public may find it difficult to score outcomes because they may misunderstand the terminology. Hence, we developed a simple questionnaire with understandable language for patients. There were 43 outcomes/outcome domains in the questionnaire. The list of outcomes is in Supplement 3. Patients were recruited by frontline clinicians in our team on 12 and 13 March 2020. Finally, 10 cured patients agreed to participate in the survey. They were asked to choose which outcomes were important to them. The characteristics of patients are shown in Table 5. More than 50% patients care about outcomes of Pulmonary imaging, lung function, respiratory symptoms such as cough and dyspnea, fever, SARS-CoV-2 nucleic acid tests, recovery, and mental.

**Table 5.** The characteristics of the patients in the survey

| Characteristics | No. of population | Characteristics | No. of population |
| --- | --- | --- | --- |
| Gender |  | Type of disease |  |
| Male | 6 (6/10, 60%) | Mild | 4 (4/10, 40%) |
| Female | 4 (4/10, 40%) | Ordinary | 4 (4/10, 40%) |
| Age |  | Severe | 0 |
| ≤18 | 0 | Critical | 2 (2/10, 20%) |
| 18-29 | 3 (3/10, 30%) | Type of therapy |  |
| 30-39 | 5 (5/10, 50%) | TCM | 0 |
| 40-49 | 1 (1/10, 10%) | Integrated TCM and Western medicine | 9 (9/10, 90%) |
| 50-59 | 1 (1/10, 10%) | Western medicine | 1 (1/10, 10%) |

### 3.4 Consensus meeting

The consensus meeting was held on 18 March 2020 and was a video conference. Six frontline clinicians (one from a WM hospital and five from a TCM hospital) as well as one frontline nurse, one methodologist and one researcher who participated in the design of clinical trials of COVID-19 were invited to attend the consensus meeting. The participants were from Shanghai (one), Beijing (five), Tianjin (two) and Guizhou (one) and all were voting participants. All clinicians and nurses had worked in Hubei Province after the COVID-19 outbreak. Two additional participants (one coordinator and one staff member from the Chinese Clinical Trial Registry) attended the meeting but did not participate in the discussion or voting.

After reporting the results of the Delphi survey and patients’ survey, participants discussed some outcomes they believed should/should not be measured in clinical trials. After discussion, voting participants were invited to vote on which outcomes should be included in the COS of COVID-19. The outcomes voted by ≥70% of participants were included in the COS. The voting results are shown in Supplement 4. The COS of COVID-19 is shown in Table 6.

**Table 6.**
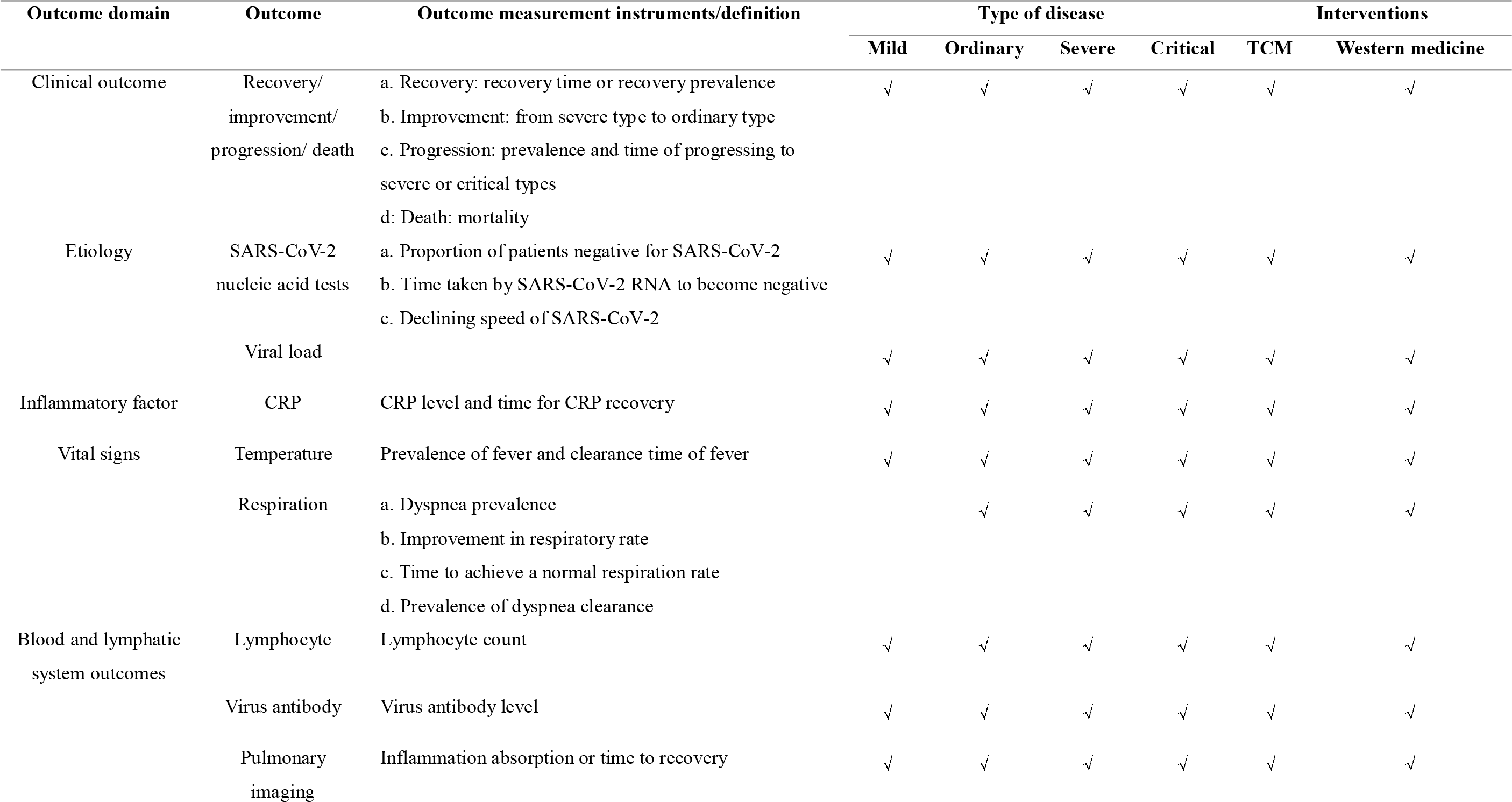
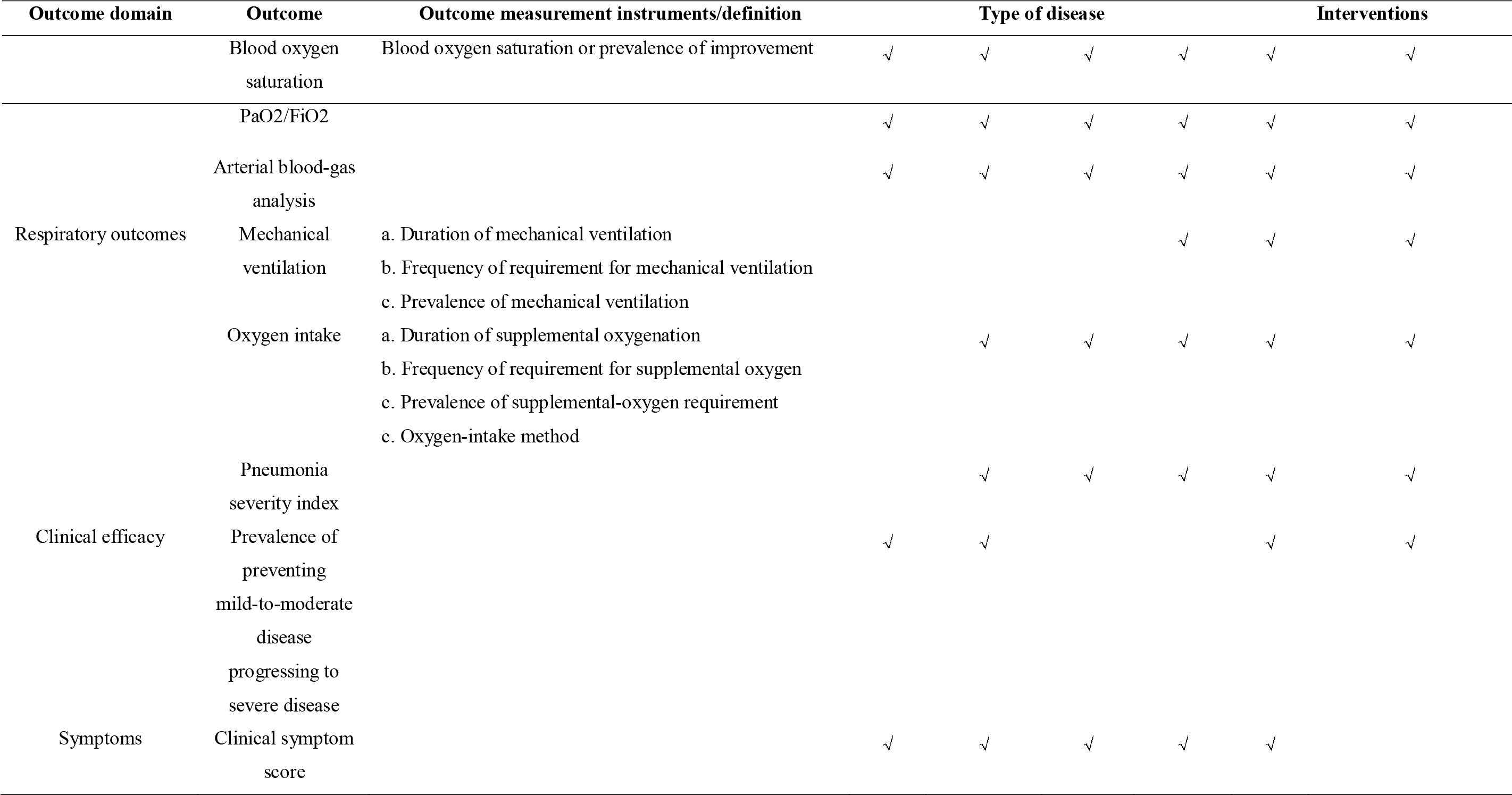
COS for COVID-19

## 4. DISCUSSION

This COS was conducted rapidly and rigorously to report an emergency in a specific environment. It can be used for any type of disease, intervention, and design. There is a specific outcome for TCM clinical trials: clinical symptom score. Researchers can measure the clinical symptom score according to different TCM syndromes. For some individuals, there are several measurements because there is no evidence to show which measurement is the best one. We hope researchers of clinical trials can use this COS to reduce heterogeneity in outcome reporting. Furthermore, our COS may help decision-makers to approve new agents for COVID-19 if researchers report important outcomes. However, researchers can report other outcomes according to the purpose of their research.

Our study had four main limitations. First, due to the highly infectious nature of SARS-CoV-2, patients and the public did not participate in the design or development of the preliminary list of outcomes. Second, the preliminary list of outcomes was developed from protocols of clinical trials when there were knowledge gaps in the prevalence, therapy, prognosis, clinical characteristics of COVID-19. Hence, the COS must be updated in the future. Third, the number of patients was small and all of them were from Hubei Province, so their perspectives may not reflect those of other regions in China or overseas. Fourth, almost all stakeholders were from China. Though one participant in round 1 of the Delphi survey was from Canada, his/her opinion reflected a Chinese perspective because the questionnaire was written in Chinese.

## Data Availability

The data is from public database and does not include identifiable patient data.

## Acknowledgements

The study team would like to thank all people who contributed to the consensus meeting and to those who took part in the Delphi survey. Those acknowledged provided permission to be mentioned as participants in the development of the COS of COVID-19.

## Delphi participants

Xuan Zhang, Jihong Feng, Yuanhao Wu, Zhong Liu, Xuezeng Hao, Dahua Wu, Yuanming Ba, Puhua Zeng, Zuomei He, Xin Yu (Beijing), Yun Li, Qinghui Zhang, Kun Cao, Jin Fang, Jun Li, Qian Li, Li Xu, Yaozu Xiang, Yonggui Wu, Song Shi, Lin Zhang, Xin Yu (Wuhan), Chao Yang, Jia He, Hong Zhang, Jingwei Zhou, Jiang Chen, Xue Li, Zhen Wang, Sushan Cao, Wenjing Liu, Yan Liu, Bo Li, Hong Zhang, Zhipeng Kang.

## Author Approval

All authors read and approved the final manuscript.

## Funding

This work was supported by the National High-level Personnel of Special Support Program (W02020052).

## Competing interests

The authors declare that there is no conflict of interest.

## Data sharing statement

The data is from public database and does not include identifiable patient data.

## Notes

### Competing Interest Statement

The authors have declared no competing interest.

### Clinical Trial

http://www.comet-initiative.org/Studies/Details/1507

### Clinical Protocols

http://www.comet-initiative.org/Studies/Details/1507

